# Estimating the contribution of HIV-infected adults to household pneumococcal transmission in South Africa, 2016-2018: A hidden Markov modelling study

**DOI:** 10.1101/2021.05.21.21257622

**Authors:** Deus Thindwa, Nicole Wolter, Amy Pinsent, Maimuna Carrim, John Ojal, Stefano Tempia, Jocelyn Moyes, Meredith McMorrow, Jackie Kleynhans, Anne von Gottberg, Neil French, Cheryl Cohen, Stefan Flasche, on behalf of PHIRST group

**Author notes:** These authors contributed equally (Cheryl Cohen, Stefan Flasche). Membership of the PHIRST group is provided in the Acknowledgements.

## Abstract

Human immunodeficiency virus (HIV) infected adults are at a higher risk of pneumococcal colonisation and disease, even while receiving antiretroviral therapy (ART). To help evaluate potential indirect effects of vaccination of HIV-infected adults, we assessed whether HIV-infected adults disproportionately contribute to household transmission of pneumococci. We constructed a hidden Markov model to capture the dynamics of pneumococcal carriage acquisition and clearance observed during a longitudinal household-based nasopharyngeal swabbing study, while accounting for sample misclassifications. Households were followed-up twice weekly for 10 months for nasopharyngeal carriage detection via real-time PCR. We estimated the effect of participant’s age, HIV status, presence of a HIV-infected adult within the household and other covariates on pneumococcal acquisition and clearance probabilities. Of 1,684 individuals enrolled, 279 (16.6%) were younger children (<5 years-old) of whom 4 (1.5%) were HIV-infected and 726 (43.1%) were adults (≥18 years-old) of whom 214 (30.4%) were HIV-infected, most (173, 81.2%) with high CD4+ count. The observed range of pneumococcal carriage prevalence across visits was substantially higher in younger children (56.9-80.5%) than older children (5-17 years-old) (31.7-50.0%) or adults (11.5-23.5%). We estimate that 14.4% (95% Confidence Interval [CI]: 13.7-15.0) of pneumococcal-negative swabs were false negatives. Daily carriage acquisition probabilities among HIV-uninfected younger children were similar in households with and without HIV-infected adults (hazard ratio: 0.95, 95%CI: 0.91-1.01). Longer average carriage duration (11.4 days, 95%CI: 10.2-12.8 vs 6.0 days, 95%CI: 5.6 - 6.3) and higher median carriage density (622 genome equivalents per millilitre, 95%CI: 507-714 vs 389, 95%CI: 311.1-435.5) were estimated in HIV-infected vs HIV-uninfected adults. The use of ART and antibiotics substantially reduced carriage duration in all age groups, and acquisition rates increased with household size. Although South African HIV-infected adults on ART have longer carriage duration and density than their HIV-uninfected counterparts, they show similar patterns of pneumococcal acquisition and onward transmission.

**Author summary:** We assessed the contribution of HIV-infected adults to household pneumococcal transmission by applying a hidden Markov model to pneumococcal cohort data comprising 115,595 nasopharyngeal samples from 1,684 individuals in rural and urban settings in South Africa. We estimated 14.4% of sample misclassifications (false negatives), representing 85.6% sensitivity of a test that was used to detect pneumococcus. Pneumococcal carriage prevalence and acquisition rates, and average duration were usually higher in younger or older children than adults. The use of ART and antibiotics reduced the average carriage duration across all age and HIV groups, and carriage acquisition risks increased in larger household sizes. Despite the longer average carriage duration and higher median carriage density in HIV-infected than HIV-uninfected adults, we found similar carriage acquisition and onward transmission risks in the dual groups. These findings suggest that vaccinating HIV-infected adults on ART with PCV would reduce their risk for pneumococcal disease but may add little to the indirect protection against carriage of the rest of the population.

## Introduction

*Streptococcus pneumoniae* (pneumococcus) caused an estimated 3.7 million cases of invasive pneumococcal disease (IPD) and 317,300 deaths in children <5 years-old, globally in 2015 [1,2]. While severe disease is largely concentrated in young children and older adults, human immunodeficiency virus (HIV)-infected adults are also at an increased risk of both colonisation and IPD [3–7]. HIV affects the T and B cell function, resulting in impaired responses to control pneumococcal carriage at mucosal level [8–10]. Although the universal scale-up of antiretroviral therapy (ART) [11,12] has successfully reduced IPD risk in HIV-infected adults [13,14], the IPD risk remains elevated if compared to HIV-uninfected adults [5,6]. ART partially reconstitutes mucosal immunity by increasing B and T cell quantity and functionality [8,15], but deficiencies in humoral mucosal response due to depleted or persistent defects in memory cell function persist after ART initiation [16–18].

Despite mature pneumococcal conjugate vaccine (PCV) infant immunisation programmes, continued circulation of vaccine preventable serotypes in adults has been observed throughout Africa [19–25]. In some countries, such as Malawi, Mozambique, and Kenya, this intersects with areas of high HIV prevalence. Adult HIV prevalence in Africa remains high [26] as a consequence of improved survival with ART use [11,12] and persistently high HIV incidence [27], thus the high risk of pneumococcal carriage and IPD in HIV-infected adults in Africa remains a concern.

Vaccination of African HIV-infected adults with PCV, similar to the recommendations in many high-income countries, may not only reduce their disease burden but also vaccine serotype pneumococcal acquisition and hence onward transmission and may thus benefit non-vaccinated populations [28]. We hypothesised that children living with HIV-infected adults have higher rates of pneumococcal carriage acquisition due to increased exposure from frequently colonised HIV-infected adults who usually have a prolonged higher carriage prevalence [5]. In this study, we assessed whether HIV-infected adults contribute more to pneumococcal transmission within the household than their HIV-uninfected counterparts.

## Methods

### Data description

The temporal dynamics of pneumococcal colonisation were observed in a cohort study (Prospective Household Observational Cohort Study of Influenza, Respiratory Syncytial Virus and Other Respiratory Pathogens Community Burden and Transmission Dynamics - PHIRST) conducted between 2016 and 2018 in a rural (Agincourt) and an urban (Klerksdorp) community in South Africa. Households were randomly selected, and were eligible for the study if they had ≥3 household members and the household members resided in the household for ≥1 year prior to study commencement, had no plan to relocate during study duration, and consented to participate in the study. Also, enrolment ensured that more than half of the households included at least one child aged <5 years, and a new cohort was enrolled every study year [29,30].

A total of 1,684 individuals from 327 households were enrolled and followed up from May to October in 2016 and January to October in 2017 and 2018. The median household size was 5 (interquartile range 4-7). Nasopharyngeal (NP) swabs were taken twice weekly, resulting in 115,595 total NP samples from 1,684 individuals [28]. The swabs were tested for the presence of pneumococci using real-time quantitative polymerase chain reaction (qPCR), targeting the autolysin (*lytA*) gene [31]. Serotyping was not performed. On enrolment, all household members were tested for HIV infection and the demographic characteristics of the study participants was recorded.

### Modelling framework

We used a continuous time, time homogeneous, hidden Markov model (HMM) which assumed a Susceptible – Infected – Susceptible (SIS) framework [32–38], to fit to individual level trajectories of colonisation during the study period. An individual can be either infected (I) and currently carrying pneumococci or be susceptible (S). Thus, the model can be described by transition intensities between S and I for acquisition (*q*_12_) and clearance (*q*_21_) in the transition intensity matrix 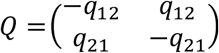.Covariates for acquisition and clearance rates are incorporated via a proportional hazard. To obtain the transition probabilities, matrix 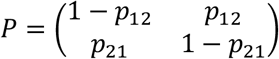 is defined and explicitly calculated through matrix exponential, *P* = exp(*Q*(*t*)), where *p*_12_ is the probability of being in state 2 (*I*) at time *t* > 0, given that the previous state was 1 (*S*). A more detailed description of the Markov transition process is provided in the Supplement.

In the hidden Markov modelling (HMM) framework [34,39–44], the states *S* and *I* of the Markov Chain (*X*_*i*_(*t*)) for individual *i* at time *t* are not observed directly, but approximated by the results of a NP swab. The link between the modelled, true infection status and observed pneumococcal carriage states in the model (*Y*_*i*_(*t*)) is governed by emission probabilities conditional on the unobserved state. We assumed 100% specificity of the NP swab and the PCR (no false positive) while estimating the proportion of false negative results (*e*) probabilistically (observed vs hidden/truth states). Hence, the emission matrix is given as 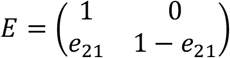 where e_21_ = Pr(*Y*_*i*_(*t*) = 1 | *X*_*i*_(*t*) = 2).

We assumed that the observed states are conditionally independent given the values of the unobserved states and that the future Markov chain is independent of its history beyond the current state (Markov property) (Fig 1). Thus, the likelihood is the product of the emission probability density and the transition probability of hidden Markov chain summed over all possible paths of the hidden states (explicitly defined in the Supplement).

**Figure 1.**
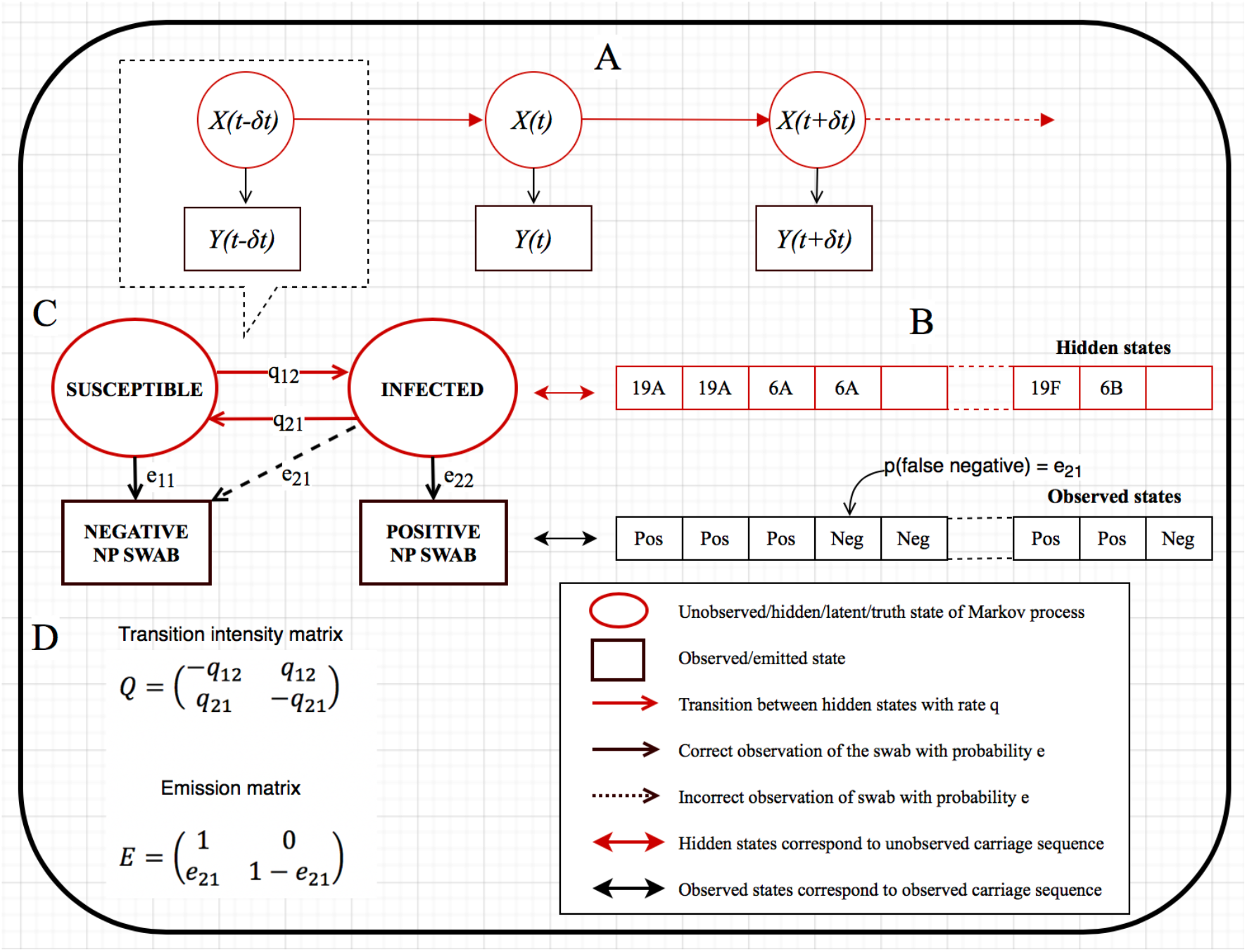
Susceptible-infected-susceptible (SIS) hidden Markov model schemas of pneumococcal carriage dynamics in South African households between 2016-2018. Continuous-time time-homogeneous hidden Markov model where *X*(*t*) represents hidden states, and *Y*(*t*) observed states, and in which *Y*(*t*) is conditionally independent given *X*(*t*) and the Markov property holds (A). Pneumococcal nasopharynx (NP) carriage sequence of a specified individual representing hidden and observed states, with a probability that an individual truly carrying a pneumococcal serotype may be detected negative by a real-time quantitative polymerase chain reaction test (B). An SIS hidden Markov model structure that captures a snapshot of part A and carriage sequence of part B in order to estimate transition rates and probability of misclassification/false negative (C). Transition intensity matrix, Q, and emission matrix, E, respectively capture the SIS model transition rates and emission or misclassification probability in part C to compute the maximum likelihood estimates of transition intensities and misclassification probability (D).

Our model assumed that carriage acquisition at the current observation point was a function of individual age group (younger child aged <5 years, older child aged 5-17 years, or adult aged ≥18 years), HIV status (infected or uninfected), number of HIV-infected adult(s) in the household, place of carriage exposure (household or community), and household size. Carriage duration was modified by individual age, HIV status, ART status, and antibiotic use. The place of carriage exposure is generally unknown without fine-scale serotype data. Crudely, we assumed that if a household member is currently infected while all other household members were susceptible at the last observation point, then current carriage acquisition of that member was attributable to community transmission [34]. Otherwise we assumed that the transmission was from within the same household (Fig 1).

### Model fit, convergence and prediction

The model was fitted to longitudinal data of pneumococcal carriage dynamics in a maximum likelihood framework using Bound Optimisation By Quadratic Approximation optim algorithm facilitated by msm R package [34,45]. To ascertain convergence of the model, we purposefully selected five unique pairs of initial transition intensities {S, I} for the Q matrix, then refitted the model five times, each time starting a Markov chain with a unique dyad and iterating 1,000 times to obtain similar final transition intensities and -2log-likelihood. Model predictions were assessed by comparing infection and susceptibility prevalence in 14-day intervals for the observed data to the fitted values. (S1 Fig) [34].

### Decoding the underlying carriage sequence

After fitting the HMM, a Viterbi algorithm with the msm function was used to recursively construct the sequence of pneumococcal carriage with the highest probability through the hidden states [46]. The probability of each hidden state at each observation point, conditionally on all the data was computed using Baum-Welch forward/backward algorithm. Thus, an overall misclassification probability of the observed states given the hidden states was computed. Model estimates of carriage transition intensity and probability were adjusted for misclassification probability (S2 Fig).

### Sensitivity analysis

In a sensitivity analysis, three alternative and potentially more parsimonious models were fitted separately to the data. Fits of these models were compared to the main model using the on Akaike Information Criterion (AIC) [47] and checked whether they yielded qualitatively different results to the main model. Each of the four fitted models assumed the same number of covariates to modify carriage acquisition intensity but varying number of covariates assumed to modify carriage duration. Potential modifiers of carriage duration included age and HIV status for model 1; age, HIV status and antibiotic use for model 2; age, HIV status and viral load based ART status for model 3; and age, HIV status, antibiotic use and viral load based ART status for main model 4 (S1 Table).

Further, we examined the impact of alternative stratification of covariates on the changes in carriage transition probabilities: (i) while the main analysis estimated age- and HIV-stratified carriage acquisition rates comparing households with ≥1 HIV-infected adult(s) versus households without HIV-infected adults, in the sensitivity analysis, we estimated age- and HIV-stratified carriage acquisition rates comparing households with 0,1, 2, 3, 4 and 5 HIV-infected adult(s) and (ii) rather than assuming time-homogeneous intensities throughout the study period, we relaxed this assumption by fitting a time-inhomogeneous model with piecewise follow-up periods; 0-99 days, 100-139, 140-199, 200-219 and 220-289 (S3 Fig).

Statistical significance was set at <0.05. All analyses were conducted in R v3.5.0 [34,48] and are available via https://github.com/deusthindwa/hmm.pneumococcus.hiv.south-africa.

### Ethical approval

The longitudinal pneumococcal carriage data described in this study were obtained from consenting South African children and adults as part of the PHIRST study. The use of data was granted by the University of Witwatersrand, Human Research Ethics Committee (HREC) and the Protocol Review Committee (PRC) under approval #150808, the US CDC’s Institutional Review Board relied on the local review (#6840), and the London School of Hygiene & Tropical Medicine Observational Research Ethics Committee under approval #17902.

## Results

### Descriptive analysis

A total of 327 households were recruited in the PHIRST study of which 166 (50.8%) had at least one member living with HIV infection. At enrolment, of 1,684 individuals included in the study, 279 (16.6%) were younger children aged less than 5 years old of whom 4 (1.5%) were HIV-infected, and 679 (40.3%) were older children aged between 5-17 years old of whom 31 (4.7%) were HIV-infected. Among the 726 (43.1%) study participants aged 18 years or older (“adults”), 214 (30.4%) were HIV-infected, and 505 (69.6%) were females. Among those HIV-infected adults, 196 (86.7%) self-reported to be on ART, although only 151 (79.5%) had CD4+ cell count of more than 350. Most adults were non-smokers (69.6%) and did not regularly consume alcohol (57.4%). A similar proportion of children lived in households with (50.6%) and without (49.4%) at least one HIV-infected adult. Among 231 younger children with vaccine information available, 227 (98.3%) received first PCV dose at 6 weeks, 225 (97.4%) second dose at 14 weeks and 216 (93.5%) third dose at 9 months of age (Table 1).

**Table 1.** Baseline demographic and clinical characteristics of children and adults who were followed up twice weekly for ten months for nasopharynx swabbing for pneumococcal carriage in South African households between 2016 and 2018.

| Description | Total<br>N=1,684 | Younger children<br><5 years<br>n=279 (16.6%) | Older children<br>5-17 years<br>n=679 (40.3%) | Adults<br>≥18 years<br>n=726 (43.1%) |
| --- | --- | --- | --- | --- |
| Mean age (SD) | 22.1 (±19.8) | 2.2 (±1.3) | 10.5 (±3.7) | 40.5 (±16.7) |
| Study site |  |  |  |  |
| Agincourt (rural) | 849 (50.4) | 171 (61.3) | 376 (55.4) | 302 (41.6) |
| Klerksdorp (semi-urban) | 835 (49.6) | 108 (38.7) | 303 (44.6) | 424 (58.4) |
| Sex |  |  |  |  |
| Female | 1,009 (59.9) | 137 (49.1) | 367 (54.1) | 505 (69.6) |
| Male | 675 (40.1) | 142 (50.9) | 312 (45.9) | 221 (30.4) |
| HIV status |  |  |  |  |
| Negative (-) | 1,379 (84.7) | 256 (98.5) | 634 (95.3) | 489 (69.6) |
| Positive (+) | 249 (15.3) | 4 (1.5) | 31 (4.7) | 214 (30.4) |
| Viral-load based ART status<sup>†</sup> |  |  |  |  |
| Not on ART | 93 (40.8) | 1 (50.0) | 13 (52.0) | 79 (39.3) |
| On ART | 135 (59.2) | 1 (50.0) | 12 (48.0) | 122 (60.7) |
| Self-reported ART status |  |  |  |  |
| Not on ART | 30 (13.2) | 0 (0.0) | 1 (3.8) | 29 (14.5) |
| On ART | 198 (86.8) | 2 (100.0) | 25 (96.2) | 171 (85.5) |
| Mean CD4 count (SD) | 673 (±430) | 1,210 (±13.4) | 857 (±418) | 645 (±426) |
| CD4+ cell count<sup>‡</sup> |  |  |  |  |
| Low | 49 (22.8) | 0 (0.0) | 10 (43.5) | 39 (20.5) |
| High | 166 (77.2) | 2 (100.0) | 13 (56.5) | 151 (79.5) |
| Living with ≥1 HIV+ adults |  |  |  |  |
| No | 818 (48.7) | 133 (47.8) | 346 (51.1) | 339 (46.9) |
| Yes | 860 (51.3) | 145 (52.2) | 331 (48.9) | 384 (53.1) |
| PCV13 doses received<sup>#</sup> |  |  |  |  |
| At 6 weeks | 227 (98.3) | 227 (98.3) | N/A | N/A |
| At 14 weeks | 225 (97.4) | 225 (97.4) | N/A | N/A |
| At 9 months | 216 (93.5) | 216 (93.5) | N/A | N/A |
| Smoking (≥18 years) |  |  |  |  |
| No | 505 (69.6) | N/A | N/A | 505 (69.6) |
| Yes | 221 (30.4) | N/A | N/A | 221 (30.4) |
| Alcohol use (≥18 years) |  |  |  |  |
| No | 417 (57.4) | N/A | N/A | 417 (57.4) |
| Yes | 309 (42.6) | N/A | N/A | 309 (42.6) |

<sup>†</sup> ART use status based on viral load results (on ART = Undetected; Not on ART = <20 copies per ml)
<sup>‡</sup> Low CD4+ count ≤350 cells/mm<sup>3</sup> and high CD4+ count >350 cells/mm<sup>3</sup> in adults, and low CD4+ count ≤750 cells/mm<sup>3</sup> and high CD4+ count >750 cells/mm<sup>3</sup> in children, in HIV-INFECTED only
<sup>#</sup> Pneumococcal conjugate vaccine (PCV13) vaccination status in younger children from available records
Standard deviation (SD)
N/A not applicable

### Carriage prevalence and density

Among HIV-uninfected participants, observed pneumococcal carriage prevalence was higher in younger children (range across visits: 56.9-80.5%) than older children (31.7-50.0%) and was lowest in adults (11.5-23.5%) (Fig 2A). Among HIV-infected participants, pneumococcal carriage prevalence fluctuated in younger children (0-100%), in older children (30-77%), and in adults (14-34%) (Fig 2A). The likelihood of detecting pneumococcal carriage during visits was higher for children than adults and for HIV-infected younger children or older children or adults than their HIV-uninfected counterparts (Fig 2B). Carriage prevalence among younger HIV-uninfected children was lower in households with less than 6 members (65.5%, 95%CI: 64.5-66.5) than in households with 6-10 (72.5%, 95%CI: 71.5-73.5) or household more than 10 members (85.6%, 95%CI: 82.4-88.8) but it was similar in HIV-infected children across household size groups (Fig 2C). Carriage prevalence fluctuated across visits by HIV-infection and sex in adults, with similar ranges between HIV-uninfected male adults 10.8-25.3% and HIV-uninfected female adults 10.2-24.0%, and between HIV-infected male adults 6.3-40.7% and HIV-infected female adults 12.3-34.6% (S5A Fig).

**Figure 2.**
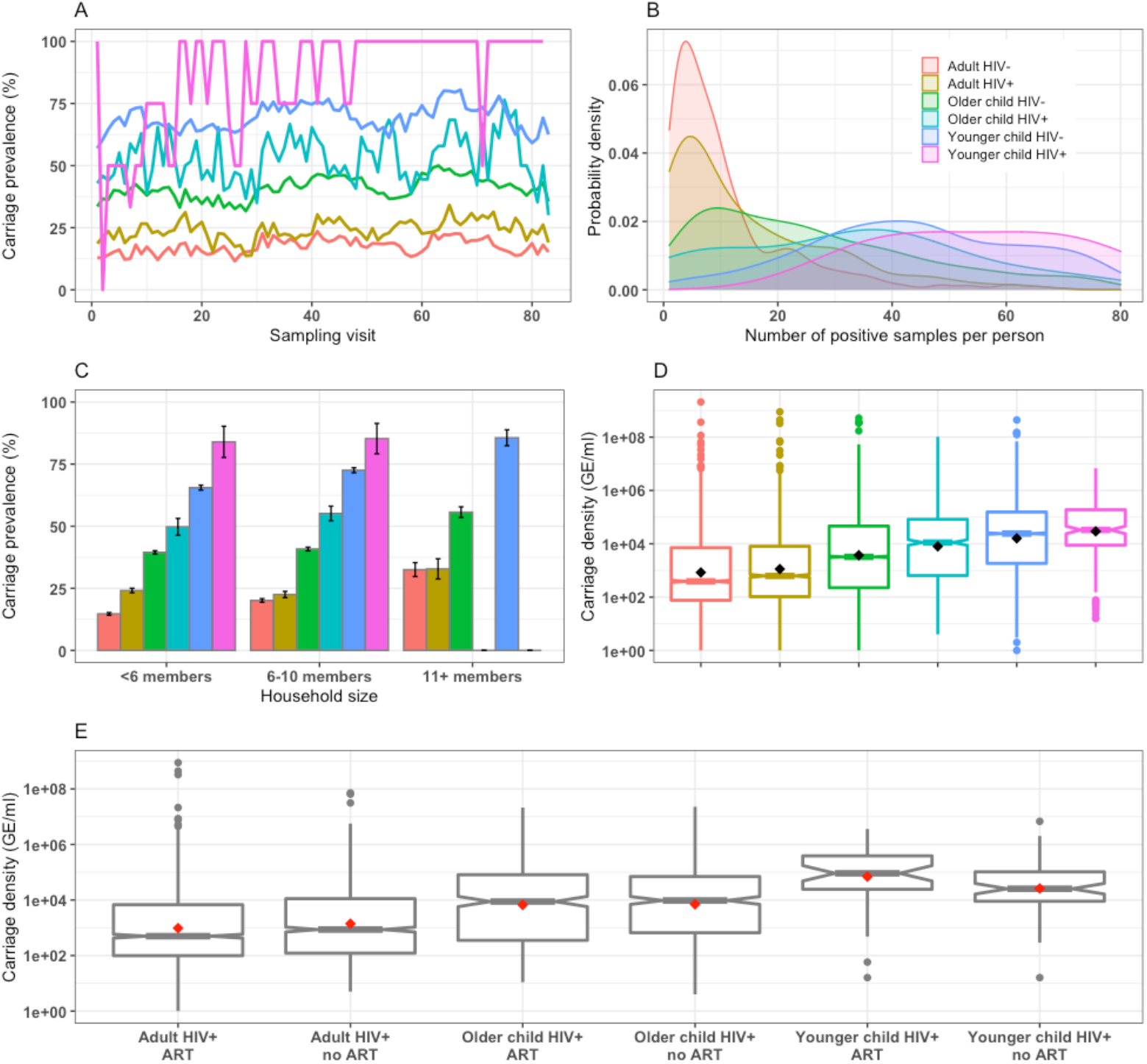
Human immunodeficiency virus (HIV)-stratified pneumococcal carriage dynamics in younger children (<5 years-old), older children (5-17 years-old) and adults (≥18 years-old) in South African households between 2016-2018. Age and HIV-stratified pneumococcal carriage prevalence by different nasopharyngeal sampling visits (A), the likelihood of detecting pneumococcal carriage during visits (B), pneumococcal carriage prevalence by household sizes with 95% confidence intervals (CI) (C) and carriage densities with mean (black diamond), median and associated 95%CI of median, 25^th^ and 75^th^ percentiles, minimum and maximum, and outlier where carriage density is measured as genome equivalents per millilitre (GE/ml) (D). Age and antiretroviral therapy (ART) stratified carriage density with mean (red diamonds), median and associated 95%CI of median, 25^th^ and 75^th^ percentiles, minimum and maximum, and outlier (notched boxplot) (E)

Median pneumococcal carriage density, in genome equivalents per millilitre (GE/ml), was significantly higher in younger children (24,341, 95%CI: 22,638-26,122) than older children (3,490, 95%CI: 3,168-3,754) or adults (476, 95%CI: 429-522). Also, median carriage density was higher in HIV-infected than HIV-uninfected older children (11,156, 95%CI: 8,681-13,948 vs 3,221, 95%CI: 2,911-3,472) or adults (622, 95%CI: 507-714 vs 389, 95%CI: 311-435), and not in younger children (33,050, 95%CI: 22,690-42,293 vs 24,124, 95%CI: 22,547-25,838) (Fig 2D). Conversely, median carriage density was similar between those not on ART compared to those on ART in older children (9,624, 95%CI: 5,289-11,843 vs 8,818, 95%CI: 5,102-12,720) or adults (861, 95%CI: 508-1,001 vs 499, 95%CI: 382-586), and not in younger children (25,430, 95%CI: 13,138-40,245 vs 91,566, 95%CI: 43,355-265,628) (Fig 2E). About 14.4%, 95%CI: 13.7-15.0 of negative NP swab results were estimated probabilistically to be false negatives.

### Pneumococcal carriage acquisition

Overall, pneumococcal carriage acquisition was higher in older children (1.15, 95%CI: 1.08-1.23) and younger children (1.52, 95%CI: 1.38-1.68) than adults. Acquisition of carriage was more frequently observed when at least another household member was infected half a week before (and hence attributed to household transmission) than in previously uninfected households (1.80, 95%CI: 1.68-1.93). Irrespective of age and HIV status, acquisition rates from within the household increased with household size; by 1.05 (95%CI: 1.00-1.10) in households with 6-10 members and by 1.41 (95%CI: 1.24-1.60) in households with 11 or more members compared to households with less than 6 members. However, within household carriage acquisition rates in children, irrespective of age group and HIV status, were not higher in the households with at least one HIV-infected adult (0.95, 95%CI: 0.91-1.01) (Table 2, Fig 3). In addition, daily carriage acquisition rates in HIV-uninfected younger children did not significantly vary between households with HIV-infected female adults (0.14, 95%CI: 0.12-0.17) and those with HIV-infected male adults (0.13, 95%CI: 0.11-0.15) (S5 Fig).

**Table 2.** Effects of included covariates on pneumococcal acquisition and clearance rates estimated in the hidden Markov model.

| Description | Hazard Ratio (95%CI)<sup>‡</sup> |
| --- | --- |
| <u>Pneumococcal carriage acquisition</u> |  |
| Age in years (y) |  |
| Adult, ≥18y | Reference |
| Older child, 5-17y | 1.15 (1.08-1.23) |
| Younger child, <5y | 1.52 (1.38-1.68) |
| HIV status |  |
| Negative | Reference |
| Positive | 0.95 (0.87- 1.04) |
| Children living with ≥1 HIV-infected adults |  |
| No | Reference |
| Yes | 0.95 (0.91- 1.01) |
| Place of carriage exposure |  |
| Community | Reference |
| Within household | 1.80 (1.68- 1.93) |
| Household size |  |
| <6 members | Reference |
| 6-10 members | 1.05 (1.00-1.10) |
| 11+ members | 1.41 (1.24-1.60) |
| <u>Pneumococcal carriage clearance</u> |  |
| Age in years (y) |  |
| Adult, ≥18y | Reference |
| Older child, 5-17 | 0.34 (0.31-0.36) |
| Younger child, <5y | 0.10 (0.09-0.12) |
| HIV status |  |
| Negative | Reference |
| Positive | 0.52 (0.46-0.59) |
| Antibiotic use |  |
| No | Reference |
| Yes | 1.47 (0.67-3.25) |
| Viral load-based ART status<sup>†</sup> |  |
| Not on ART | Reference |
| On ART | 1.29 (1.13-1.47) |
| <sup>†</sup> ART use status based on viral load results<br><sup>‡</sup> 95% confidence interval (95%CI) |  |

**Figure 3.**
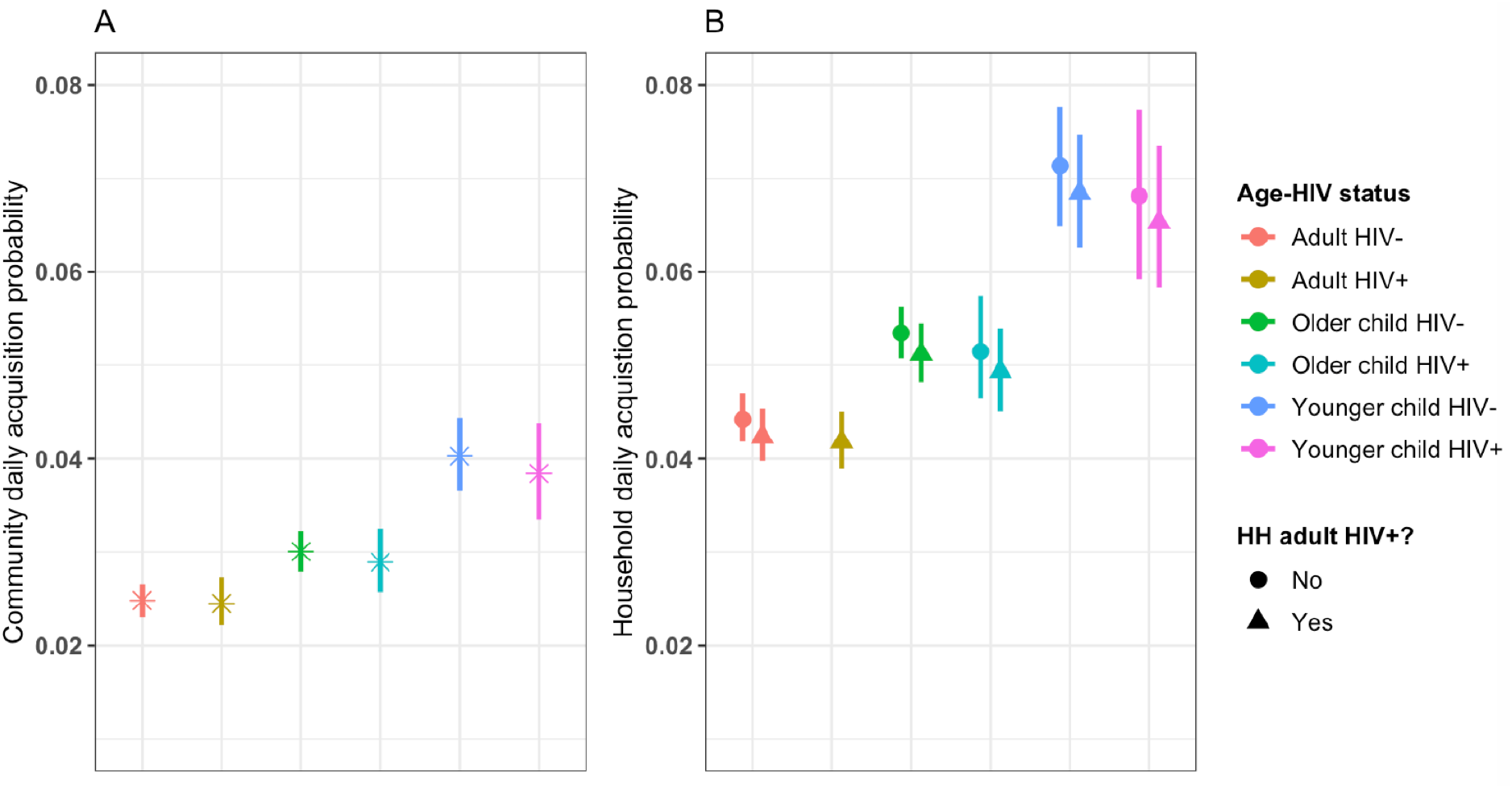
Community and within household (HH) acquisitions of pneumococcal carriage in younger children (<5 years-old), older children (5-17 years-old) and adults (≥18 years-old) in South African households between 2016-2018. Age and human immunodeficiency virus (HIV) stratified estimates of community carriage acquisition probability per day (A) and within household carriage acquisition probability per day over the total follow-up period (B), comparing household without HIV-infected adult(s) (HIV-) to households with HIV-infected adult(s) (+).

We estimated 3.8 carriage acquisition episodes per year, (95%CI: 3.4-4.2) for HIV-infected younger children, 5.9 (95%CI: 5.4-6.3) for HIV-uninfected younger children, 7.4 (95%CI: 6.7-8.1) for HIV-infected older children and 10.6 (95%CI: 10.2-11.0) for HIV-uninfected older children from households with at least one HIV-infected adult, and these were similar to their counterparts from households without HIV-infected adults (3.8, 95%CI: 3.3-4.2 and 5.8, 95%CI: 5.4-6.3, and 7.3, 95%CI: 6.6-8.0 and 10.3, 95%CI: 9.9-10.8, respectively) (Fig 3, S2 Table).

### Pneumococcal carriage duration

The average duration of pneumococcal carriage was highest in HIV-infected and HIV-uninfected younger children (107.9 days, 95%CI: 92.1-124.7 and 56.3 days, 95%CI: 51.1-62.1) followed by HIV-infected and HIV-uninfected older children (33.9 days, 95%CI: 29.9-38.6 and 17.9 days, 95%CI: 16.8-18.5), and HIV-infected and HIV-uninfected adults (11.4 days, 95%CI: 10.2-12.8 and 6.0 days, 95%CI: 5.6-6.3) (Fig 4C, Fig 4D, and S3 Table).

**Figure 4.**
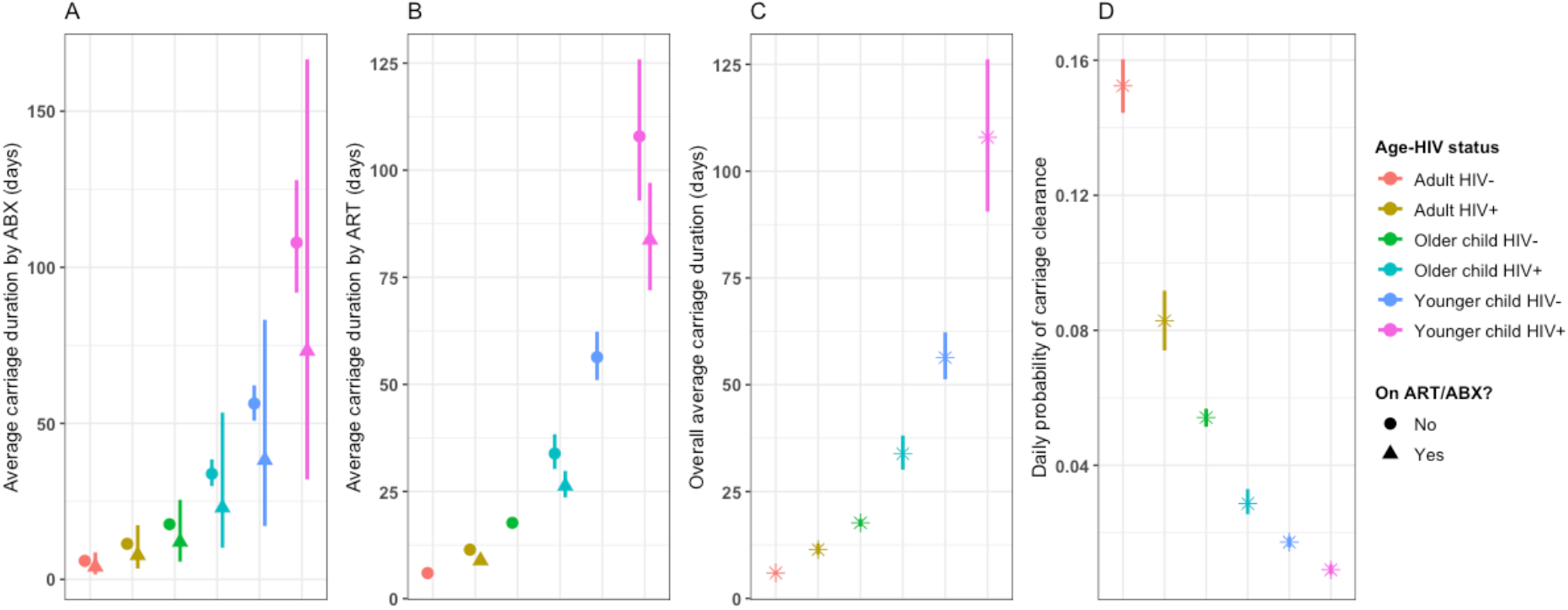
Duration of pneumococcal carriage in younger children (<5 years-old), older children (5-17 years-old) and adults (≥18 years-old) in South African households between 2016-2018. Age and human immunodeficiency virus (HIV) stratified average carriage duration in days comparing individuals on antibiotics (ABX) (triangular shape) to those not on ABX (circular shape) (A), and individuals on antiretroviral therapy (ART) (triangle shape) to those not on ART (circle shape) (B). Age and HIV stratified overall average carriage duration in days (C). Age and HIV stratified daily probability of carriage clearance (D).

Pneumococcal carriage cleared slower in older children (Hazard Ratio [HR]: 0.34, 95%CI: 0.31-0.36) and younger children (HR: 0.10, 95%CI: 0.09-0.12) when compared to adults. Carriage clearance was slower in HIV-infected than in HIV-uninfected individuals (HR: 0.52, 95%CI: 0.46-0.59), and faster in HIV-infected individuals with successfully suppressed viral load than in those without successful viral suppression (HR: 1.29, 95%CI: 1.13-1.47) (Fig 4B, Table 2). Antibiotic use may have accelerated pneumococcal clearance; however, the effect was not statistically significant (HR: 1.47, 95%CI: 0.67-3.25) (Fig 4A, Table 2).

### Sensitivity analysis

In the sensitivity analysis, a model that included age, HIV status, antibiotic use, and ART status as potential modifiers for pneumococcal carriage duration had the lowest AIC score as well as for including both antibiotic use and ART status (S1 Table). Increasing the number of HIV-infected adults within household to 1, 2, 3, 4, and 5 resulted in similar estimates of pneumococcal carriage acquisition in younger or older children (S3A Fig). Our results were also robust when instead of assuming a time homogeneous hidden Markov model, we allowed for the estimation of time varying transition probabilities (S3B Fig)

## Discussion

We used a HMM to better understand pneumococcal carriage dynamics, and the role of HIV-infected adults in it, using data from a densely sampled longitudinal South African cohort using data from 115,595 nasopharyngeal swabs. We estimated that children have higher acquisition rates and duration of carriage than adults, and that, within a household, HIV-infected adults are not more likely to transmit pneumococci to children than HIV-uninfected adults. Pneumococcal acquisition events increased with larger household size irrespective of age and HIV status. Although ART use reduced pneumococcal carriage duration in HIV-infected children and adults, they still carry pneumococci for longer than their HIV-uninfected counterparts.

Higher household acquisition rates in children than adults have been reported previously [32,49–53], although not consistently across studies [35,54]. This may reflect setting-specific population mixing behaviour and immunisation levels. Similarly, and for the first time in the presence of a mature infant PCV routine vaccination programme, we find that children both have higher acquisition rates than adults and carry pneumococci for longer, making them a likely key source for pneumococcal transmission in and beyond the household [55,56].

We postulated that HIV-infected adults were more likely to carry pneumococci and may have higher carriage density which individually or in combination may increase their risk for pneumococcal transmission compared to HIV-uninfected adults. Prior to infant PCV introduction, a study in Malawi showed that HIV-infected adults on ART had higher carriage prevalence than those not on ART [5], and two studies in South Africa also found that HIV-infected adults (mothers) had higher carriage prevalence than their HIV-uninfected counterparts, irrespective of ART status [32,49]. In addition, HIV-infected adults (mothers) were found to transmit pneumococci to their children more often than HIV-uninfected peers [32]. We generated additional evidence showing that, in the PCV era, carriage prevalence is slightly increased in HIV-infected adults on ART compared to HIV-uninfected adults as a result of reduced carriage clearance rates. We also show that median carriage density is higher in HIV-infected than HIV-uninfected adults. However, we find no evidence that carriage density is modified by ART status in HIV-infected adults (Fig 2). Further research may need to investigate whether differential effects of ART on pneumococcal carriage density in adults by country may be driven by types of ART regimens used.

Furthermore, our model estimates that the presence of an HIV-infected adult in the household does not increase the risk for pneumococcal carriage acquisition in co-habiting children. Although it is possible that there may have been other HIV-infected adults within households who were not enrolled into the study, it is unlikely this would alter the results given the insensitive acquisition estimates with increasing number of HIV-infected adults within household (S3A Fig). These findings support the notion that ART largely, but not completely, reconstitutes the anti-pneumococcal mucosal immune response in HIV-infected adults [8]. This would imply that HIV-infected adults do not contribute disproportionally to pneumococcal transmission when on ART and hence that their vaccination is unlikely to substantially add to the herd protection already induced by the childhood immunisation programme although vaccination will provide direct protection against IPD in HIV-infected adults.

Our observation of increasing pneumococcal carriage acquisition rates with higher household size has also been reported previously [36] and suggests density dependent transmission in the household [57]. In line with evidence before infant PCV introduction [32,36], we find that pneumococcal carriage acquisition probabilities from the community were higher in children than in adults irrespective of HIV status, likely in part due to frequent effective contacts among playschool children [49,58] and immature immunity in children relatively to adults. We also estimate that children were twice as likely to get infected from within the household than from the community. However, we base this inference on the identified pneumococcal carriage in a household member at the previous visit.

In the absence of serotyping of the pneumococcal isolates, our inferences may be prone to overestimation within household transmission by linking family members who in fact were infected with different pneumococcal serotypes. Similarly, serotyping would enhance our ability to differentiate a single and long carriage episode from almost immediate re-acquisition or the clearance of the dominant serotype while the previously subdominant serotype persists. This may have led to an overestimation of carriage duration and underestimated clearance rates. However, the mean carriage duration of 56 days (51-62) in HIV-uninfected children estimated in this study aligns with studies that used serotype data [32,35,49,59]. While both the estimates for duration of carriage and the contribution of household transmission may be somewhat exaggerated, the lack of serotyping should not have affected our primary outcome, the relative contribution of HIV infected adults to pneumococcal transmission.

The use of ART, as inferred from measured viral load in study participants, reduced pneumococcal carriage duration by 22% compared to no ART use within each age group of HIV-infected participants. However, mean pneumococcal carriage duration remained slightly higher than their HIV-uninfected counterparts (Fig 4). Our model also estimated the sensitivity of the swabbing and qPCR testing regime for the detection of pneumococcal carriage. We estimate that about 1 in 7 swabs were misclassified as pneumococcal negative. False negatives might have arisen as a result of the sampling technique or if samples contained insufficient quantities of bacteria to successfully amplify and detect [58]. We assumed 100% specificity of an assay targeting the autolysin gene as the probability of false positives is seemingly very low [31,59]. Our estimated misclassification probability in this study is within 10-20% range of values that were reported elsewhere [59,60].

In conclusion, we used one of the most densely sampled longitudinal pneumococcal carriage studies to infer the role of HIV-infected adults in pneumococcal transmission in the PCV and ART era. We find that the transmission risk from HIV-infected adults largely aligns with that of their uninfected counterpart. This implies that PCV use in HIV-infected adults who have access to ART would reduce their risk for pneumococcal disease but may have little added benefit over vaccinating other adults to the indirect protection against carriage of the rest of the population.

## Data Availability

Data cannot be shared publicly because of confidentiality. Data are available from the
National Institute of Communicable Disease (NICD) if authorised by Institutional Data
Access / Ethics Committee (contact via Professor Cherly Cohen)
for researchers who meet the criteria for access to confidential data.
The code underlying the results presented in the study are available from GitHub
through the following link
(https://github.com/deusthindwa/hmm.pneumococcus.hiv.south-africa) or contact Deus
Thindwa through

## Acknowledgement

DT, OJ are supported by the National Institute for Health Research (NIHR) Global Health Research Unit on Mucosal Pathogens (MPRU) using UK aid from the UK Government (16/136/46). AP is supported by the Bill and Melinda Gates Foundation. SF is supported by a Sir Henry Dale Fellowship jointly funded by the Wellcome Trust and the Royal Society (Grant number 208812/Z/17/Z). CC and AvG receive grant support through their institution from Sanofi Pasteur. The funders had no involvement in the study design; collection, analysis and interpretation of data; writing of the report; or decision to submit the article for publication. Members of the Prospective Household Observational Cohort Study of Influenza, Respiratory Syncytial Virus and Other Respiratory Pathogens Community Burden and Transmission Dynamics (PHIRST) have played a role of funding-acquisition and administration. The following are members of the PHIRST group: Centre for Respiratory Diseases and Meningitis, National Institute for Communicable Diseases of the National Health Laboratory Service, Johannesburg, South Africa (Amelia Buys, Lorens Maake, Florette Treurnicht, Orienka Hellferscee, Thulisa Mkhencele); Environment and Health Research Unit, South African Medical Research Council, Johannesburg, South Africa (Angie Mathee); Unit for Environmental Science and Management, School of Geo- and Spatial Science, North-West University, Potchefstroom, South Africa (Brigitte Language, Stuart Piketh); School of Public Health, Faculty of Health Sciences, University of the Witwatersrand, Johannesburg (Lorens Maake, Floidy Wafawanaka, Ryan G Wagner, F. Xavier Gómes-Olivé, Kathleen Kahn); Perinatal HIV Research Unit, MRC Soweto Matlosana Collaborating Centre for HIV/AIDS and TB, University of the Witwatersrand, Johannesburg, South Africa (Katlego Mothlaoleng, Limakatso Lebina, Neil A Martinson); Johns Hopkins University Center for TB Research, Baltimore, Maryland, United States of America (Neil A Martinson). We are grateful to all individuals who participated in the PHIRST study. We also thank all community leaders from Agincourt Klerksdorp for allowing us to conduct PHIRST study in these areas.

## Author contribution

Conceptualization: DT, SF, NF; Methodology: DT, SF, AP; Writing-Original Draft: DT; Writing-Review and Editing: DT, NW, AP, CM, JO, ST, JM, MM, JK, AvG, NF, CC, SF; Funding-Acquisition: DT, CC, AvG. All authors read and approved the final manuscript.

## Disclaimer

The findings and conclusions in this publication are those of the authors and do not necessarily represent the official position of the United States Centers for Disease Control and Prevention, the NIHR or the Department of Health and Social Care.

## Reference

1. O’Brien KL, Wolfson LJ, Watt JP, Henkle E, Deloria-Knoll M, McCall N, et al. Burden of disease caused by Streptococcus pneumoniae in children younger than 5 years: global estimates. The Lancet. 2009;374: 893–902. doi:10.1016/S0140-6736(09)61204-6

2. Wahl B, O’Brien KL, Greenbaum A, Majumder A, Liu L, Chu Y, et al. Burden of Streptococcus pneumoniae and Haemophilus influenzae type b disease in children in the era of conjugate vaccines: global, regional, and national estimates for 2000–15. The Lancet Global Health. 2018;6: e744–e757. doi:10.1016/S2214-109X(18)30247-X

3. van Aalst M, Lötsch F, Spijker R, van der Meer JTM, Langendam MW, Goorhuis A, et al. Incidence of invasive pneumococcal disease in immunocompromised patients: A systematic review and meta-analysis. Travel Medicine and Infectious Disease. 2018;24: 89–100. doi:10.1016/j.tmaid.2018.05.016

4. Corcoran M, Vickers I, Mereckiene J, Murchan S, Cotter S, Fitzgerald M, et al. The epidemiology of invasive pneumococcal disease in older adults in the post-PCV era. Has there been a herd effect? Epidemiology & Infection. 2017;145: 2390–2399. doi:10.1017/S0950268817001194

5. Heinsbroek E, Tafatatha T, Phiri A, Ngwira B, Crampin A, Read J, et al. Persisting high prevalence of pneumococcal carriage among HIV-infected adults receiving antiretroviral therapy in Malawi. Aids. 2015;29: 1837–1844. doi:10.1097/QAD.0000000000000755

6. Nunes MC, Gottberg A von, Gouveia L de, Cohen C, Kuwanda L, Karstaedt AS, et al. Persistent High Burden of Invasive Pneumococcal Disease in South African HIV-Infected Adults in the Era of an Antiretroviral Treatment Program. PLOS ONE. 2011;6: e27929. doi:10.1371/journal.pone.0027929

7. Gill CJ, Mwanakasale V, Fox MP, Chilengi R, Tembo M, Nsofwa M, et al. Impact of Human Immunodeficiency Virus Infection on Streptococcus pneumoniae Colonization and Seroepidemiology among Zambian Women. J Infect Dis. Oxford Academic; 2008;197: 1000–1005. doi:10.1086/528806

8. Zhang L, Li Z, Wan Z, Kilby A, Kilby JM, Jiang W. Humoral immune responses to Streptococcus pneumoniae in the setting of HIV-1 infection. Vaccine. 2015;33: 4430–4436. doi:10.1016/j.vaccine.2015.06.077

9. Jochems SP, Weiser JN, Malley R, Ferreira DM. The immunological mechanisms that control pneumococcal carriage. PLOS Pathogens. 2017;13: e1006665. doi:10.1371/journal.ppat.1006665

10. Iwajomo OH, Finn A, Ogunniyi AD, Williams NA, Heyderman RS. Impairment of Pneumococcal Antigen Specific Isotype-Switched Igg Memory B-Cell Immunity in HIV Infected Malawian Adults. PLOS ONE. 2013;8: e78592. doi:10.1371/journal.pone.0078592

11. Harries AD, Ford N, Jahn A, Schouten EJ, Libamba E, Chimbwandira F, et al. Act local, think global: how the Malawi experience of scaling up antiretroviral treatment has informed global policy. BMC Public Health. 2016;16: 938. doi:10.1186/s12889-016-3620-x

12. Jahn A, Harries AD, Schouten EJ, Libamba E, Ford N, Maher D, et al. Scaling-up antiretroviral therapy in Malawi. Bulletin of the World Health Organization. 2016;94: 772–776. doi:10.2471/BLT.15.166074

13. Heffernan RT, Barrett NL, Gallagher KM, Hadler JL, Harrison LH, Reingold AL, et al. Declining Incidence of Invasive Streptococcus pneumoniae Infections among Persons with AIDS in an Era of Highly Active Antiretroviral Therapy, 1995—2000. J Infect Dis. 2005;191: 2038–2045. doi:10.1086/430356

14. Everett DB, Mukaka M, Denis B, Gordon SB, Carrol ED, Oosterhout JJ van, et al. Ten Years of Surveillance for Invasive Streptococcus pneumoniae during the Era of Antiretroviral Scale-Up and Cotrimoxazole Prophylaxis in Malawi. PLOS ONE. 2011;6: e17765. doi:10.1371/journal.pone.0017765

15. Nunes MC, Madhi SA. Safety, immunogenicity and efficacy of pneumococcal conjugate vaccine in HIV-infected individuals. Human Vaccines & Immunotherapeutics. 2012;8: 161–173. doi:10.4161/hv.18432

16. Eley B. Immunization in Patients with HIV Infection. Drugs. 2008;68: 1473–1481. doi:10.2165/00003495-200868110-00001

17. De Milito A, Mörch C, Sönnerborg A, Chiodi F. Loss of memory (CD27) B lymphocytes in HIV-1 infection. AIDS. 2001;15: 957.

18. Madhi SA, Kuwanda L, Cutland C, Holm A, Käyhty H, Klugman KP. Quantitative and Qualitative Antibody Response to Pneumococcal Conjugate Vaccine Among African Human Immunodeficiency Virus-Infected and Uninfected Children. The Pediatric Infectious Disease Journal. 2005;24: 410. doi:10.1097/01.inf.0000160942.84169.14

19. Swarthout TD, Fronterre C, Lourenço J, Obolski U, Gori A, Bar-Zeev N, et al. High residual carriage of vaccine-serotype Streptococcus pneumoniae after introduction of pneumococcal conjugate vaccine in Malawi. Nature Communications. Nature Publishing Group; 2020;11: 2222. doi:10.1038/s41467-020-15786-9

20. Lourenço J, Obolski U, Swarthout TD, Gori A, Bar-Zeev N, Everett D, et al. Determinants of high residual post-PCV13 pneumococcal vaccine-type carriage in Blantyre, Malawi: a modelling study. BMC Medicine. 2019;17. doi:10.1186/s12916-019-1450-2

21. Heinsbroek E, Tafatatha T, Phiri A, Swarthout TD, Alaerts M, Crampin AC, et al. Pneumococcal carriage in households in Karonga District, Malawi, before and after introduction of 13-valent pneumococcal conjugate vaccination. Vaccine. 2018;36: 7369–7376. doi:10.1016/j.vaccine.2018.10.021

22. Hammitt LL, Etyang AO, Morpeth SC, Ojal J, Mutuku A, Mturi N, et al. Effect of ten-valent pneumococcal conjugate vaccine on invasive pneumococcal disease and nasopharyngeal carriage in Kenya: a longitudinal surveillance study. The Lancet. 2019; doi:10.1016/S0140-6736(18)33005-8

23. Sigaúque B, Moiane B, Massora S, Pimenta F, Verani JR, Mucavele H, et al. Early Declines in Vaccine Type Pneumococcal Carriage in Children Less Than 5 Years Old After Introduction of 10-valent Pneumococcal Conjugate Vaccine in Mozambique. The Pediatric Infectious Disease Journal. 2018;37: 1054. doi:10.1097/INF.0000000000002134

24. Usuf E, Bottomley C, Bojang E, Cox I, Bojang A, Gladstone R, et al. Persistence of Nasopharyngeal Pneumococcal Vaccine Serotypes and Increase of Nonvaccine Serotypes Among Vaccinated Infants and Their Mothers 5 Years After Introduction of Pneumococcal Conjugate Vaccine 13 in The Gambia. Clin Infect Dis. 2019;68: 1512–1521. doi:10.1093/cid/ciy726

25. Klugman KP, Rodgers GL. Population versus individual protection by pneumococcal conjugate vaccination. The Lancet. 2019;393: 2102–2104. doi:10.1016/S0140-6736(19)30039-X

26. Dwyer-Lindgren L, Cork MA, Sligar A, Steuben KM, Wilson KF, Provost NR, et al. Mapping HIV prevalence in sub-Saharan Africa between 2000 and 2017. Nature. 2019;570: 189. doi:10.1038/s41586-019-1200-9

27. UNAIDS. Global HIV & AIDS databook [Internet]. Geneva, Switzerland: Joint United Nations Programme on HIV/AIDS; 2017 pp. 1–248. Available: https://www.unaids.org/sites/default/files/media_asset/20170720_Data_book_2017_en.pdf

28. Thindwa D, Pinsent A, Ojal J, Gallagher KE, French N, Flasche S. Vaccine strategies to reduce the burden of pneumococcal disease in HIV-infected adults in Africa. Expert Review of Vaccines. Taylor & Francis; 2020;0: 1–8. doi:10.1080/14760584.2020.1843435

29. Cohen C, McMorrow M, Martinson NA, Kahn K, Treurnicht FK, Moyes J, et al. Cohort Profile: a Prospective Household cohort study of Influenza, Respiratory Syncytial virus, and other respiratory pathogens community burden and Transmission dynamics in South Africa (PHIRST), 2016-2018. medRxiv. Cold Spring Harbor Laboratory Press; 2021; 2021.01.06.21249313. doi:10.1101/2021.01.06.21249313

30. Cohen C, Kleynhans J, Moyes J, McMorrow ML, Treurnicht FK, Hellferscee O, et al. Asymptomatic transmission and high community burden of seasonal influenza in an urban and a rural community in South Africa, 2017–18 (PHIRST): a population cohort study. The Lancet Global Health. 2021;9: e863–e874. doi:10.1016/S2214-109X(21)00141-8

31. Blaschke AJ. Interpreting Assays for the Detection of Streptococcus pneumoniae. Clin Infect Dis. Oxford Academic; 2011;52: S331–S337. doi:10.1093/cid/cir048

32. Shiri T, Auranen K, Nunes MC, Adrian PV, van Niekerk N, de Gouveia L, et al. Dynamics of Pneumococcal Transmission in Vaccine-Naïve Children and Their HIV-infected or HIV-uninfected Mothers During the First 2 Years of Life. Am J Epidemiol. 2013;178: 1629–1637. doi:10.1093/aje/kwt200

33. Cox DR, Miller HD. The Theory of Stochastic Processes. CRC Press; 1977.

34. Jackson C. Multi-State Models for Panel Data: The msm Package for R. Journal of Statistical Software. 2011;38: 1–28. doi:10.18637/jss.v038.i08

35. Lipsitch M, Abdullahi O, D’Amour A, Xie W, Weinberger D, Tchetgen ET, et al. Estimating Rates of Carriage Acquisition and Clearance and Competitive Ability for Pneumococcal Serotypes in Kenya With a Markov Transition Model. Epidemiology. 2012;23: 510–519. doi:10.1097/EDE.0b013e31824f2f32

36. Melegaro A, Gay NJ, Medley GF. Estimating the transmission parameters of pneumococcal carriage in households. Epidemiol Infect. 2004;132: 433–441.

37. Melegaro A, Choi Y, Pebody R, Gay N. Pneumococcal Carriage in United Kingdom Families: Estimating Serotype-specific Transmission Parameters from Longitudinal Data. Am J Epidemiol. 2007;166: 228–235. doi:10.1093/aje/kwm076

38. Jones E, Epstein D, García-Mochón L. A Procedure for Deriving Formulas to Convert Transition Rates to Probabilities for Multistate Markov Models. Medical Decision Making. 2017;37: 779–789. doi:10.1177/0272989X17696997

39. Jackson CH, Sharples LD. Hidden Markov models for the onset and progression of bronchiolitis obliterans syndrome in lung transplant recipients. Statistics in Medicine. 2002;21: 113–128. doi:10.1002/sim.886

40. Jackson CH, Sharples LD, Thompson SG, Duffy SW, Couto E. Multistate Markov models for disease progression with classification error. Journal of the Royal Statistical Society: Series D (The Statistician). 2003;52: 193–209. doi:10.1111/1467-9884.00351

41. Bureau A, Shiboski S, Hughes JP. Applications of continuous time hidden Markov models to the study of misclassified disease outcomes. Statistics in Medicine. 2003;22: 441–462. doi:10.1002/sim.1270

42. Cooper B, Lipsitch M. The analysis of hospital infection data using hidden Markov models. Biostatistics. 2004;5: 223–237. doi:10.1093/biostatistics/5.2.223

43. Satten GA, Longini IM. Markov Chains With Measurement Error: Estimating the ‘True’ Course of a Marker of the Progression of Human Immunodeficiency Virus Disease. Journal of the Royal Statistical Society Series C (Applied Statistics). 1996;45: 275–309. doi:10.2307/2986089

44. McClintock BT, Langrock R, Gimenez O, Cam E, Borchers DL, Glennie R, et al. Uncovering ecological state dynamics with hidden Markov models. Ecology Letters. 2020;23: 1878–1903. doi:https://doi.org/10.1111/ele.13610

45. Powell MJD. The BOBYQA algorithm for bound constrained optimization without derivatives. Cambridge; 2009. p. 39. Available: http://www.damtp.cam.ac.uk/user/na/NA_papers/NA2009_06.pdf

46. Viterbi A. Error bounds for convolutional codes and an asymptotically optimum decoding algorithm. IEEE Transactions on Information Theory. 1967;13: 260–269. doi:10.1109/TIT.1967.1054010

47. Stone M. An Asymptotic Equivalence of Choice of Model by Cross-Validation and Akaike’s Criterion. Journal of the Royal Statistical Society Series B (Methodological). 1977;39: 44–47.

48. R Core Team (2018). R: A language and environment for statistical computing. R Foundation for Statistical Computing, Vienna, Austria. [Internet]. [cited 28 May 2019]. Available: https://www.r-project.org/

49. Nunes MC, Shiri T, van Niekerk N, Cutland CL, Groome MJ, Koen A, et al. Acquisition of Streptococcus pneumoniae in Pneumococcal Conjugate Vaccine-naïve South African Children and Their Mothers. The Pediatric Infectious Disease Journal. 2013;32: e192. doi:10.1097/INF.0b013e31828683a3

50. Heinsbroek E, Tafatatha T, Chisambo C, Phiri A, Mwiba O, Ngwira B, et al. Pneumococcal Acquisition Among Infants Exposed to HIV in Rural Malawi: A Longitudinal Household Study. Am J Epidemiol. 2016;183: 70–78. doi:10.1093/aje/kwv134

51. Turner P, Turner C, Jankhot A, Helen N, Lee SJ, Day NP, et al. A Longitudinal Study of Streptococcus pneumoniae Carriage in a Cohort of Infants and Their Mothers on the Thailand-Myanmar Border. PLOS ONE. Public Library of Science; 2012;7: e38271. doi:10.1371/journal.pone.0038271

52. Hussain M, Melegaro A, Pebody RG, George R, Edmunds WJ, Talukdar R, et al. A longitudinal household study of Streptococcus pneumoniae nasopharyngeal carriage in a UK setting. Epidemiology & Infection. 2005;133: 891–898. doi:10.1017/S0950268805004012

53. Althouse BM, Hammitt LL, Grant L, Wagner BG, Reid R, Larzelere-Hinton F, et al. Identifying transmission routes of Streptococcus pneumoniae and sources of acquisitions in high transmission communities. Epidemiology and Infection. 2017;145: 2750–2758. doi:10.1017/S095026881700125X

54. Abdullahi O, Karani A, Tigoi CC, Mugo D, Kungu S, Wanjiru E, et al. Rates of Acquisition and Clearance of Pneumococcal Serotypes in the Nasopharynges of Children in Kilifi District, Kenya. J Infect Dis. 2012;206: 1020–1029. doi:10.1093/infdis/jis447

55. Flasche S, Lipsitch M, Ojal J, Pinsent A. Estimating the contribution of different age strata to vaccine serotype pneumococcal transmission in the pre vaccine era: a modelling study. BMC Medicine. 2020;18. doi:10.1186/s12916-020-01601-1

56. Wyllie AL, Warren JL, Regev-Yochay G, Givon-Lavi N, Dagan R, Weinberger DM. Serotype Patterns of Pneumococcal Disease in Adults Are Correlated With Carriage Patterns in Older Children. Clinical Infectious Diseases. 2020; doi:10.1093/cid/ciaa1480

57. Endo A, Uchida M, Kucharski AJ, Funk S. Fine-scale family structure shapes influenza transmission risk in households: Insights from primary schools in Matsumoto city, 2014/15. PLOS Computational Biology. Public Library of Science; 2019;15: e1007589. doi:10.1371/journal.pcbi.1007589

58. Weinberger DM, Pitzer VE, Regev-Yochay G, Givon-Lavi N, Dagan R. Association Between the Decline in Pneumococcal Disease in Unimmunized Adults and Vaccine-Derived Protection Against Colonization in Toddlers and Preschool-Aged Children. Am J Epidemiol. 2019;188: 160–168. doi:10.1093/aje/kwy219

59. Lees JA, Croucher NJ, Goldblatt D, Nosten F, Parkhill J, Turner C, et al. Genome-wide identification of lineage and locus specific variation associated with pneumococcal carriage duration. In: eLife [Internet]. 25 Jul 2017 [cited 24 Jun 2019]. doi:10.7554/eLife.26255

60. Abdullahi O, Wanjiru E, Musyimi R, Glass N, Scott JAG. Validation of Nasopharyngeal Sampling and Culture Techniques for Detection of Streptococcus pneumoniae in Children in Kenya. Journal of Clinical Microbiology. American Society for Microbiology Journals; 2007;45: 3408–3410. doi:10.1128/JCM.01393-07

