## Supplementary materials for "Estimating the contribution of HIV-infected adults to household pneumococcal transmission in South Africa, 2016-2018: A hidden Markov modelling study"

‡ These authors contributed equally

* Corresponding author

**Multi-state Markov model of pneumococcal carriage dynamics**

A continuous-time time-homogeneous Markov model of pneumococcal carriage dynamics consists of susceptible (S) and infected (I) states, and 1,684 individuals, each of whom is in S or I state at any observation point. S or I state occupied by $i^{th}$ individual at time $t$ is denoted by $X_i(t)$, and movement of individuals between S and I states is governed by a set of transition intensities, $(q_{12})$ or $(q_{21})$ where {1,2}={S,I}, which may depend on the time and a set of potential individual covariates. The transition intensity represents instantaneous risk of moving from state S to I or I to S per unit time [1,2], e.g.

$$q_{12}(t; z) = \lim_{\delta_t \to 0} \Pr\left(X_i(t + \delta_t) = I \mid X_i(t) = S\right)/\delta_t \qquad (1)$$

Transition intensities form a 2X2 matrix, Q, whose rows sum up to 0 so that the diagonal entries are $q_{22} := -q_{21}\ or\ q_{11} := -q_{12}$. Thus, multi-state model fitting to observed individual pneumococcal carriage sequence data enables the Q matrix below to be estimated as follows.

$$Q = \begin{pmatrix} q_{11} & q_{12} \\ q_{21} & q_{22} \end{pmatrix} \rightarrow \begin{pmatrix} -q_{12} & q_{12} \\ q_{21} & -q_{21} \end{pmatrix} \qquad (2)$$

In this framework, the future state evolution of the Markov chain only depends on the current pneumococcal carriage state and not pneumococcal carriage history $(H_t)$ before time $t$. It's assumed that all individuals in S have the same probability to move to I and back regardless of $H_t$. However, this probability is modified by individual covariates, $z$, so that $q_{12}(t; z; H_t) = q_{12}(t; z)$ or $q_{21}(t; z; H_t) = q_{21}(t; z)$. Moreover, our time-homogeneous model assumes that a transition intensity is constant over follow-up time and pneumococcal carriage duration is distributed exponentially at a rate of $-q_{22}$ so that the mean pneumococcal carriage duration is $1/q_{21}$.

**Covariates of pneumococcal carriage dynamics in a multi-state model**

Pneumococcal carriage acquisition could be modified by age, human immunodeficiency virus (HIV), household adult HIV status, source of pneumococcal infection or household size. On the other hand, pneumococcal carriage duration may be altered by age, HIV status, antiretroviral therapy (ART) or antibiotic use. Proportional hazards model is used to estimate the effect $(\beta)$ of $i^{th}$ individual covariate

($z_i$) on transition intensity over all transitions ($T$) e.g. $q_{12}(z_i(t)) = q_{12}^{(0)} exp\left(\beta_{12}^T z_i(t)\right)$ and $q_{21}(z_i(t)) = q_{21}^{(0)} exp\left(\beta_{21}^T z_i(t)\right)$ for pneumococcal carriage acquisition and clearance respectively. The Q matrix with covariates is used for likelihood estimation, and is maximised over the baseline intensities $q_{12}^{(0)}$ and $q_{21}^{(0)}$, and the log-hazard ratios $\beta_{12}^T$ and $\beta_{21}^T$.

**Converting the transition intensity matrix into a transition probability matrix**

A transition probability matrix (P) is derived from Q matrix using matrix exponentiation, such that $P = \exp\big(Q(t)\big)$, and used for likelihood estimation. In time-homogeneous intensities, entry of P matrix, $p_{12}(t)$, defines the probability of being in state I at time *t*, given that the system was in state S at a previously time, and vice versa for $p_{21}(t)$ [2–4].

The transition probabilities form a 2X2 matrix, P, whose rows sum up to 1 so that the diagonal entries are $p_{22} := 1 - q_{21}\ or\ p_{11} := 1 - q_{12}$. The P matrix is thus defined as follows

$$P = \begin{pmatrix} p_{11} & p_{12} \\ p_{21} & p_{22} \end{pmatrix} \rightarrow \begin{pmatrix} 1 - p_{12} & p_{12} \\ p_{21} & 1 - p_{21} \end{pmatrix} \qquad (3)$$

**Hidden Markov model of pneumococcal carriage dynamics**

We extend the Markov model to a hidden Markov model (HMM) where S and I states are not directly observed [5–7]. Observed nasopharyngeal (NP) swab results are governed by probability distribution (emission probability) conditionally on unobserved state. The evolution of the hidden Markov chain is governed by Q matrix and the observed NP swab results are states assumed to be misclassifications of the hidden states. If $X_i(t)$ is the hidden Markov chain of pneumococcal carriage dynamics of individual $i$ at time $t$ only known through realisations of NP swab result $Y_i(t)$, then the sensitivity of the quantitative polymerase chain reaction (qPCR) test used to detect pneumococcus was measured by the probability that the observed state is equal to a hidden state when carriage is detected. Thus, misclassification probability = 1-sensitivity e.g. $e_{21} = \Pr(Y_i(t) = S \mid X_i(t) = I)$ [8,9].

Misclassification probabilities form a 2X2 matrix, E, whose rows sum up to 1 so that the diagonal entries are $e_{22} := 1 - e_{21}$ $or$ $e_{11} := 1$ since we assume 100% specificity (no false positive tests). The E matrix is thus defined as follows

$$E = \begin{pmatrix} e_{11} & e_{12} \\ e_{21} & e_{22} \end{pmatrix} \rightarrow \begin{pmatrix} 1 & 0 \\ e_{21} & 1-e_{21} \end{pmatrix} \qquad (4)$$

**Likelihood contribution from uncensored pneumococcal carriage states**

The likelihood estimation of the HMM is computed through forward algorithm based on matrix product, assuming the observed states are conditionally independent given the values of unobserved states and the future Markov chain is independent of past history. Thus, the individual ($i$) likelihood is integrated over all possible hidden states ($X_{i,j}$) at all observation points ($j$) adjusted for false negative NP swabs

$$L_{i,j} = \sum_{X_{i,1}} \Pr\left(Y_{i,1} \middle| X_{i,1}\right) \Pr\left(X_{i,1}\right) \ldots \ldots \sum_{X_{i,j}} \Pr\left(Y_{i,j} | X_{i,j}\right) \Pr\left(X_{ij} | X_{i,j-1}\right) \qquad (5)$$

**Likelihood contribution from censored pneumococcal carriage states**

Missing NP swab result is assumed to be censored at each observation point they occurred as its exact value is unknown but largely known to be either I or S. If $Y(t_{i,j+1})$ is the pneumococcal carriage censored state $Y$ of individual $i$ at observation time ($j + 1$), then individual likelihood is given by

$$L_{i,j} = \sum_{m \in \{I,S\}} \Pr\left(Y(t_{i,j})\right) | m \left(t_{i,j+1} - t_{i,j}\right) \qquad (6)$$

The maximum likelihood estimation is computed using Bound Optimization By Quadratic Approximation, a derivative-free iterative optim algorithm for finding the minimum -2log-likelihood of a HHM through construction of quadratic models by interpolation [10].

**Sensitivity on hidden Markov models (fitting and comparison)**

Four HMMs were fitted to the same observations using Akaike Information Criterion (AIC) defined by equation 7 [11]. AIC estimated a relative distance between the unknown true likelihood function of the data and the likelihood function of each HHM such that a lower AIC value was an indication that the fitted model was close to the true model. "Model 4" was selected because not only was its AIC score close to "Model 3" but also accounted for both antiretroviral therapy and antibiotics use.

$$AIC = -2L(\theta) + 2k \quad (7)$$

Where $L$ is the log-likelihood of a fitted HMM with parameter set $\theta$, and $k$ is the number of parameters in the fitted HMM.

Supplementary Table 1. Multistate model comparisons between hidden Markov models of specified degree of freedom (*df*) using Akaike Information Criterion (AIC) after fitting the models to longitudinal pneumococcal carriage data in South African households, 2016-2018.

| Model | Covariates for pneumococcal carriage acquisition | Covariates for pneumococcal carriage duration | *df* | AIC |
|---|---|---|---|---|
| 1 | Age + HIV status + household adult HIV status[†] + Carriage exposure[‡] + household size | Age + HIV status | 14 | 95604.11 |
| 2 | Age + HIV status + household adult HIV status[†] + Carriage exposure [‡] + household size | Age + HIV status + Antibiotic use | 15 | 95605.36 |
| 3 | Age + HIV status + household adult HIV status[†] + Carriage exposure [‡] + household size | Age + HIV status + ART status[#] | 15 | 95592.17 |
| 4 | Age + HIV status + household adult HIV status[†] + Carriage exposure [‡] + household size | Age + HIV status + Antibiotic use + ART status[#] | 16 | 95593.38* |

† Whether an individual is living in a household with HIV-infected adult(s) or HIV-uninfected adult(s) only.
‡ Whether carriage exposure may have come from within household or community (outside household).
### Antiretroviral (ART) status determined by viral load results.
* The main model 4 has AIC score close to model 3 but also accounts for both ART and antibiotic use.
HIV - Human immunodeficiency virus.

Supplementary Table 2. Maximum likelihood parameter estimates and 95% confidence intervals (95%CI) for acquisition probabilities within household and from community from a hidden Markov model that is fitted to pneumococcal carriage data in South African households, 2016-18.

| Age and HIV-specific estimates (per day) | Household with ≥1 HIV+ adult(s) (95%CI) | Household without HIV+ adult (95%CI) |
|---|---|---|
| Carriage acquisition within household | | |
| Younger child HIV+ | 0.065 (0.058 - 0.073) | 0.068 (0.060 - 0.078) |
| Younger child HIV- | 0.068 (0.062 - 0.075) | 0.071 (0.065 - 0.078) |
| Older child HIV+ | 0.049 (0.045 - 0.054) | 0.051 (0.046 - 0.058) |
| Older child HIV- | 0.051 (0.048 - 0.054) | 0.053 (0.051 - 0.056) |
| Adult HIV+ | 0.042 (0.039 - 0.045) | NA |
| Adult HIV- | 0.042 (0.040 - 0.045) | 0.044 (0.042 - 0.047) |
| Carriage acquisition from community | | |
| Younger child HIV+ | 0.038 (0.033 - 0.044) | |
| Younger child HIV- | 0.040 (0.037 - 0.044) | |
| Older child HIV+ | 0.029 (0.026 - 0.033) | |
| Older child HIV- | 0.030 (0.028 - 0.032) | |
| Adult HIV+ | 0.024 (0.022 - 0.027) | |
| Adult HIV- | 0.025 (0.023 - 0.027) | |

Human immunodeficiency virus infected (HIV+) or uninfected (HIV-)

Supplementary Table 3. Maximum likelihood parameter estimates and 95% confidence intervals (95%CI) for carriage duration and by antibiotic use and antiretroviral therapy (ART) from a hidden Markov model that is fitted to pneumococcal carriage data in South African households, 2016-18.

| Age and HIV-specific estimates | Estimate (95%CI) | Estimate (95%CI) |
|---|---|---|
| Carriage duration (days) | Antibiotics use | No antibiotics use |
| Younger child HIV+ | 73.2 (34.2 - 157.5) | 107.9 (92.4 - 125.8) |
| Younger child HIV- | 38.2 (18.5 - 82.8) | 56.3 (50.9 - 62.1) |
| Older child HIV+ | 23.0 (9.6 - 52.8) | 33.9 (29.6 - 38.4) |
| Older child HIV- | 12.0 (5.5 - 25.7) | 17.7 (16.8 - 18.5) |
| Adult HIV+ | 7.7 (3.4 - 17.1) | 11.4 (10.2 - 12.9) |
| Adult HIV- | 4.0 (1.8 - 8.9) | 6.0 (5.6 - 6.3) |
| Carriage duration (days) | On ART | Not on ART |
| Younger child HIV+ | 83.8 (72.3 - 95.5) | 107.9 (92.5 - 125.4) |
| Younger child HIV- | NA | 56.3 (50.9 - 62.3) |
| Older child HIV+ | 26.3 (23.4 - 29.6) | 33.9 (29.8 - 38.7) |
| Older child HIV- | NA | 17.7 (16.9 - 18.6) |
| Adult HIV+ | 8.9 (8.0 - 9.7) | 11.4 (10.1 - 12.7) |
| Adult HIV- | NA | 6.0 (5.6 - 6.3) |
| Carriage duration (days) | Overall | |
| Younger child HIV+ | 107.9 (92.1 - 124.7) | |
| Younger child HIV- | 56.3 (51.1 - 62.1) | |
| Older child HIV+ | 33.9 (29.9 - 38.6) | |
| Older child HIV- | 17.9 (16.8 - 18.5) | |
| Adult HIV+ | 11.4 (10.2 - 12.8) | |
| Adult HIV- | 6.0 (5.6 - 6.3) | |
| Carriage clearance probability per day | Overall | |
| Younger child HIV+ | 0.009 (0.008 - 0.011) | |
| Younger child HIV- | 0.017 (0.016 - 0.019) | |
| Older child HIV+ | 0.029 (0.025 - 0.032) | |
| Older child HIV- | 0.054 (0.052 - 0.057) | |
| Adult HIV+ | 0.083 (0.074 - 0.092) | |
| Adult HIV- | 0.152 (0.144 - 0.160) | |

**Checking convergence and predictions of a fitted Hidden Markov model**

Convergence of the selected HMM was assured by running 5 Markov chains, each with 1000 iterations. Each chain was a unique pair of initial transition intensities converging to similar final baseline intensities and -2log-likelihood. To show how the HMM predicted the irregularly-observed process, we compared the proportions of pneumococcal carriage observed with predicted, grouped over 14-days intervals. We assumed (a) individual's state at arbitrary time was the same as the state at their previously observation time given high frequent pneumococcal carriage sampling and (b) the Markov process began at a common time for all individuals [2]. If $n(t)$ individuals are under observation at time $t$, then the predicted number of individuals in I state at time $t$ is $n(t)P(t)_{0,I}$, where $P(t)$ is the transition probability matrix.

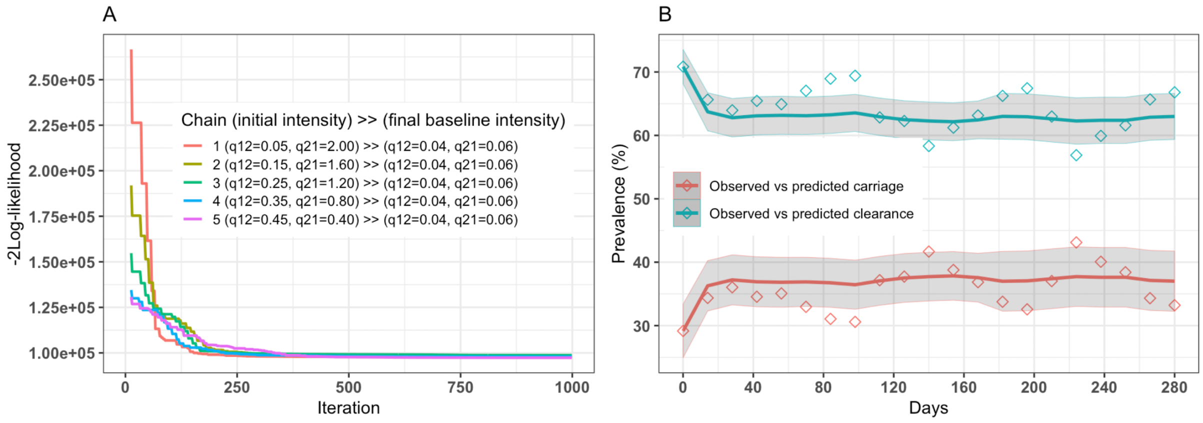

Supplementary Figure 1. Hidden Markov model (HMM) convergence and predictions. HMM convergence estimated using maximum likelihood, given 5 Markov chains each with 1000 iterations. Each chain is a unique pair of initial infected ($q_{12}$) and susceptible ($q_{21}$) intensities converging to similar final baseline transition intensities, and 2*log-likelihood (A). HMM fitting assessment comparing the observed (diamond) to predicted (line) pneumococcal carriage and clearance with 95% predictive intervals of the model-fitted line, where observed data are grouped into 14-days intervals to compute fitted values (B).

#### Viterbi algorithm

We used msm Viterbi function to recursively construct the sequence of pneumococcal carriage with the highest probability through the hidden states [12]. The probability of each hidden state at each observation point, conditionally on all the data was also computed using Baum-Welch forward/backward algorithm.

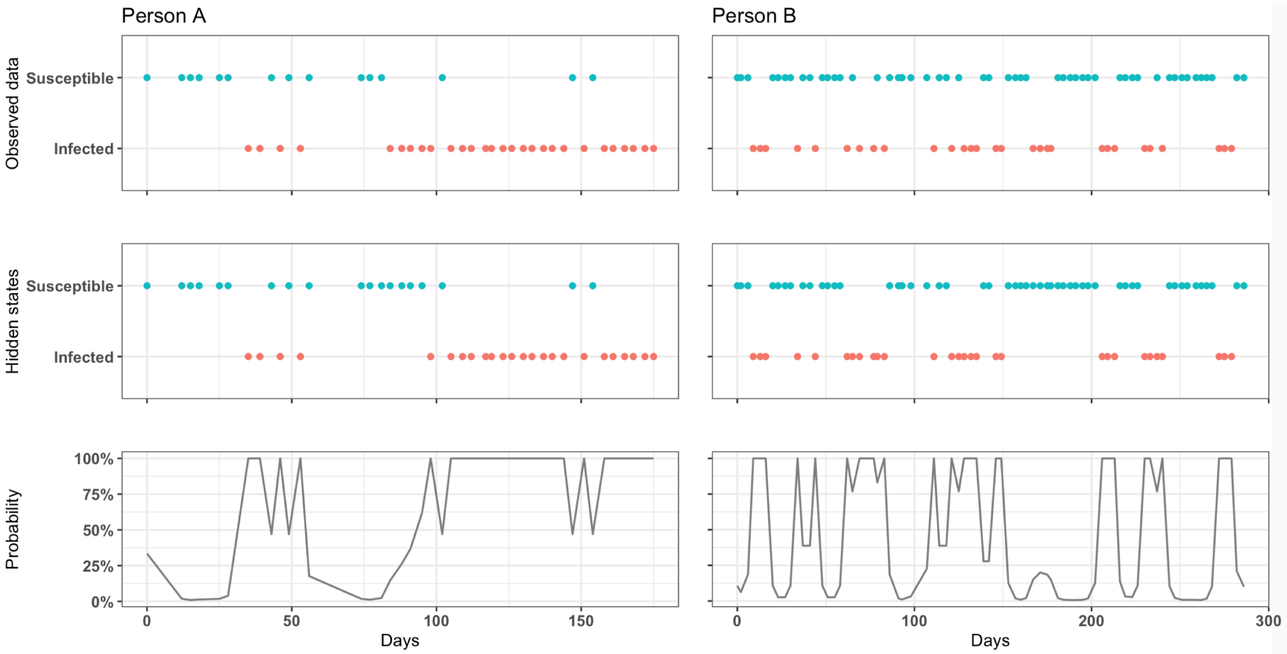

Supplementary Figure 2. The probabilities of the underlying states and the most likely path through them. Observed pneumococcal carriage results from NP swabs of two randomly selected persons A and B (first row). The underlying sequences of a fitted HMM given the observed sequence, found by the Viterbi algorithm through a recursive construction of the path with the highest probability (second row). Probability of each hidden state at each observation point, conditionally on all the data computed using Baum-Welch forward/backward algorithm (probability>50% and <100%) reflects misclassification (third row).

**Sensitivity on the number of HIV+ adults within household and time-homogeneous HMM**

We conducted a sensitivity analysis to check if varying covariate values affects pneumococcal acquisition within household. The HMM was refitted with different numbers of HIV+ adults in the household and different sampling periods.

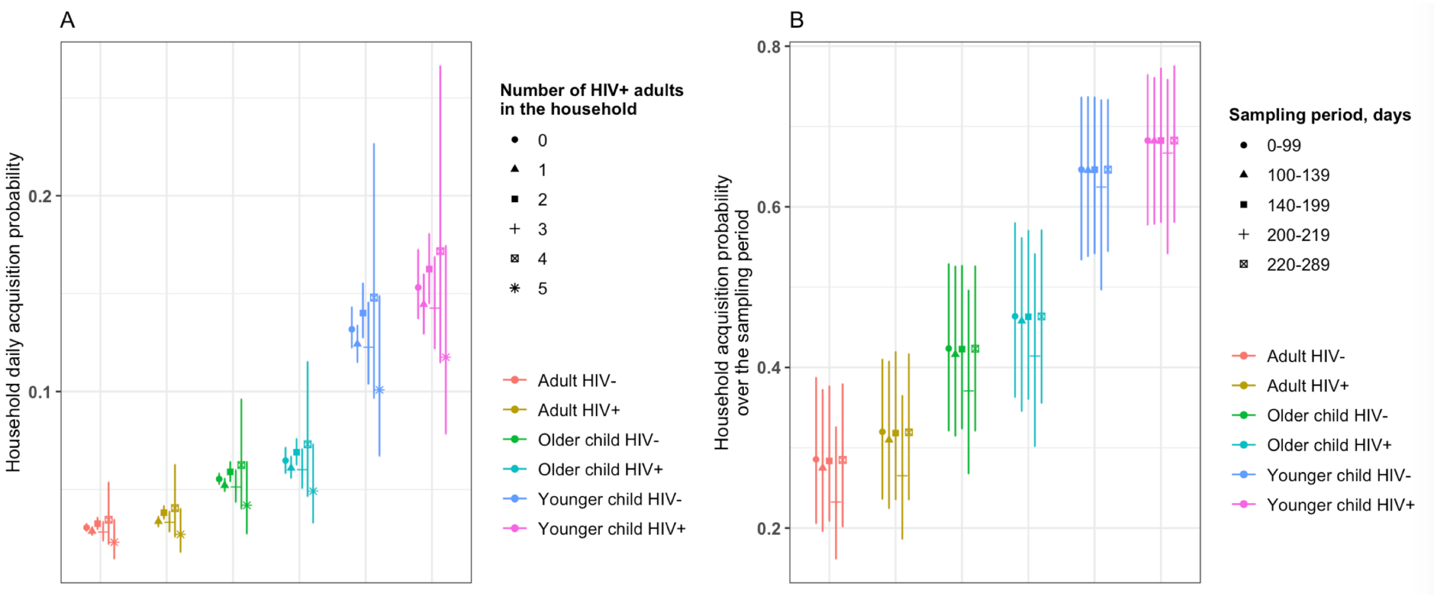

Supplementary Figure 3. Sensitivity analysis of varying covariate values in younger children (<5 years-old), older children (5-17 years-old) and adults (≥18 years-old). The main analysis computed within household HIV+ and age-stratified acquisition per day comparing households with HIV+ adult(s) to those without, whereas here, we compare households with 0,1,2,3,4 or 5 HIV+ adult(s) (A). Similarly, the main analysis estimated acquisition probabilities for entire study follow-up period (0-289 days), whereas here, we estimate acquisition probability comparing samples collected between different periods.

**HIV-age distribution, and pneumococcal carriage acquisition dynamics by household size**

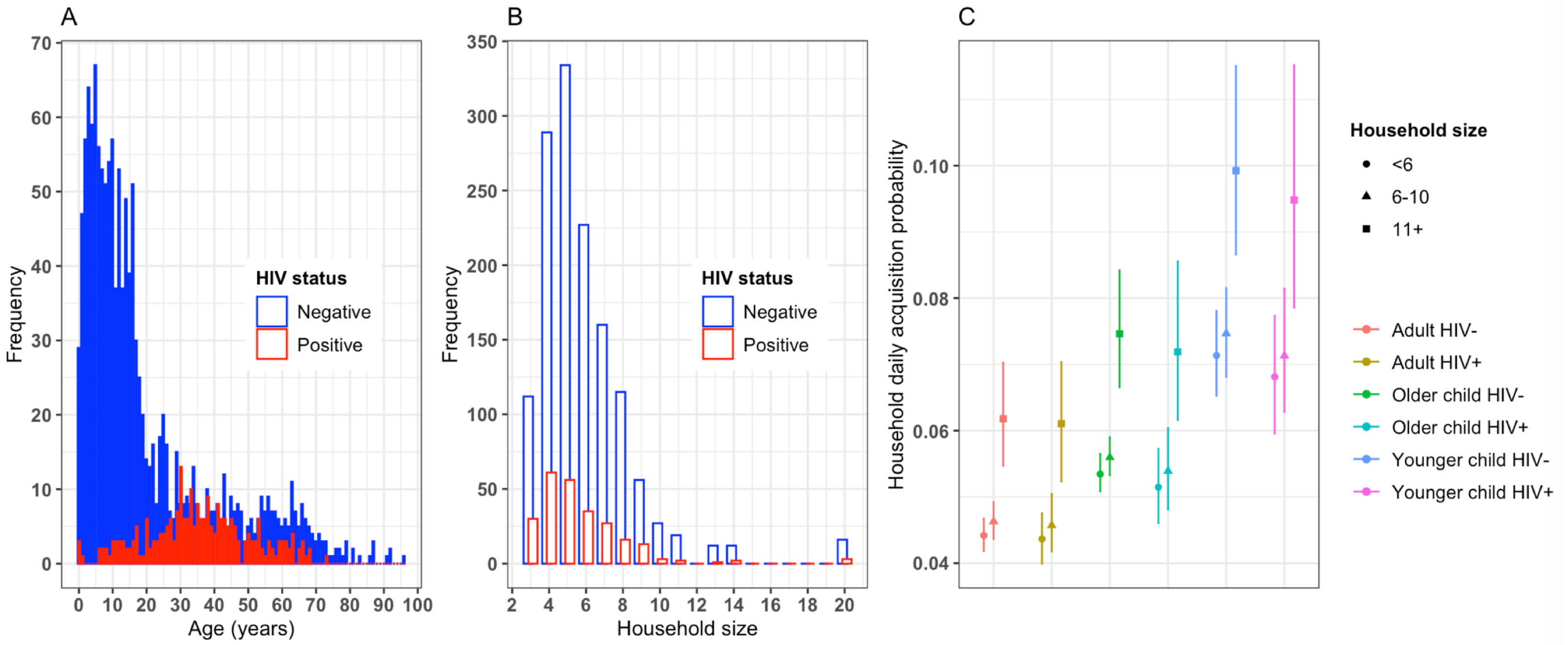

Supplementary Figure 4. HIV and Age distribution of study participants, and household size carriage acquisition dynamics in younger children (<5 years-old), older children (5-17 years-old) and adults (≥18 years-old). Overall HIV and age distribution of study participants (A). HIV and age distribution of study participants by their household size (B). HIV and age-stratified carriage acquisition probability per day by household size (C).

**Sensitivity on the contribution of HIV+ female adults to household pneumococcal transmission in children**

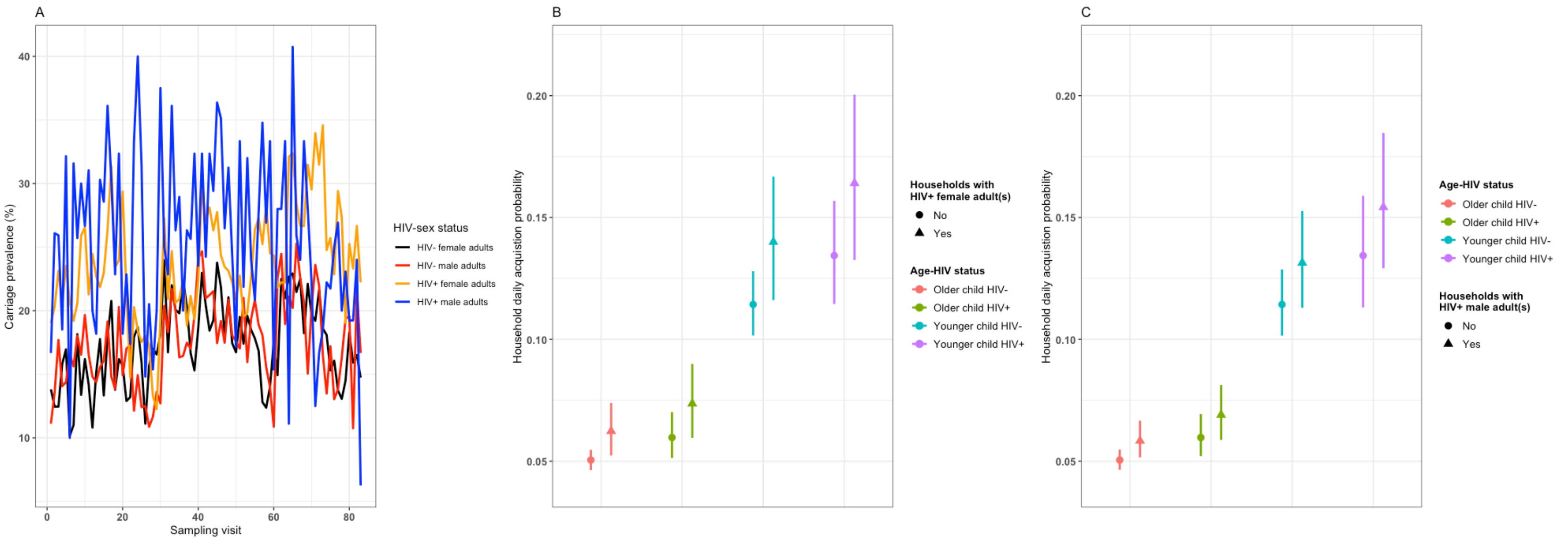

Supplementary Figure 5. Pneumococcal carriage prevalence in adults (≥18 years-old) (A), household pneumococcal acquisition in younger children (<5 years-old) and older children (5-17 years-old) by HIV status of household female adults living with these children (B), and household pneumococcal acquisition in younger and older children by HIV status of household male adults living with these children (C).

#### Reference

1. Chapman LAC, Dyson L, Courtenay O, Chowdhury R, Bern C, Medley GF, et al. Quantification of the natural history of visceral leishmaniasis and consequences for control. Parasit Vectors. 2015;8: 521. doi:10.1186/s13071-015-1136-3
2. Jackson C. Multi-State Models for Panel Data: The msm Package for R. J Stat Softw. 2011;38: 1–28. doi:10.18637/jss.v038.i08
3. Jones E, Epstein D, García-Mochón L. A Procedure for Deriving Formulas to Convert Transition Rates to Probabilities for Multistate Markov Models. Med Decis Making. 2017;37: 779–789. doi:10.1177/0272989X17696997
4. Cox DR, Miller HD. The Theory of Stochastic Processes. CRC Press; 1977.
5. Jackson CH, Sharples LD. Hidden Markov models for the onset and progression of bronchiolitis obliterans syndrome in lung transplant recipients. Stat Med. 2002;21: 113–128. doi:10.1002/sim.886
6. Bureau A, Shiboski S, Hughes JP. Applications of continuous time hidden Markov models to the study of misclassified disease outcomes. Stat Med. 2003;22: 441–462. doi:10.1002/sim.1270
7. Cooper B, Lipsitch M. The analysis of hospital infection data using hidden Markov models. Biostatistics. 2004;5: 223–237. doi:10.1093/biostatistics/5.2.223
8. Jackson CH, Sharples LD, Thompson SG, Duffy SW, Couto E. Multistate Markov models for disease progression with classification error. J R Stat Soc Ser Stat. 2003;52: 193–209. doi:10.1111/1467-9884.00351
9. Satten GA, Longini IM. Markov Chains With Measurement Error: Estimating the `True’ Course of a Marker of the Progression of Human Immunodeficiency Virus Disease. J R Stat Soc Ser C Appl Stat. 1996;45: 275–309. doi:10.2307/2986089
10. Powell MJD. The BOBYQA algorithm for bound constrained optimization without derivatives. Cambridge; 2009. p. 39. Available: http://www.damtp.cam.ac.uk/user/na/NA_papers/NA2009_06.pdf
11. Stone M. An Asymptotic Equivalence of Choice of Model by Cross-Validation and Akaike’s Criterion. J R Stat Soc Ser B Methodol. 1977;39: 44–47.
12. Viterbi A. Error bounds for convolutional codes and an asymptotically optimum decoding algorithm. IEEE Trans Inf Theory. 1967;13: 260–269. doi:10.1109/TIT.1967.1054010
